# Interventions to prevent or delay the onset of type 2 diabetes in women with prior gestational diabetes mellitus: protocol for a living systematic review and prospective meta-analysis

**DOI:** 10.1101/2021.10.31.21265595

**Authors:** Vivian Yejee Lee, Mohammad Riashad Monjur, Yashdeep Gupta, Alpesh Goyal, Gian Luca Di Tanna, Nikhil Tandon, Anushka Patel

## Abstract

**Introduction:** Gestational diabetes mellitus (GDM), once considered a transient condition during pregnancy, is now a firmly established risk factor for type 2 diabetes mellitus (T2DM). Women whose blood glucose levels do not return to normal soon after giving birth are particularly at high risk of developing established diabetes and consequent heart and blood vessel disease. Lifestyle interventions are recommended for women with GDM to prevent or delay the subsequent development of T2DM. Recent systematic reviews and meta-analyses have suggested postpartum lifestyle interventions may be beneficial in reducing the risk of developing diabetes in women with GDM, however, included studies were generally small, many had a high risk of bias and subsequent data have become available with new trials likely to complete in the next couple of years. In addition, to the best of our knowledge, formal systematic review and meta-analysis of other approaches to preventing diabetes in this population (e.g. pharmacotherapy) has not been attempted. Therefore, an updated systematic review is needed and will be formulated as a living systematic review to ensure the inclusion of emerging studies.

**Methods and analysis:** A living systematic review and a prospective meta-analysis to examine the effectiveness of postpartum interventions in reducing the risk of developing T2DM in women with recent GDM.

**Ethics and dissemination:** Ethics committee approval is not required. The data included will be from published studies, and a continued living systematic review and prospective meta-analysis will occur once a year for the next five years. Results of the review will be disseminated at relevant meetings.

**PROSPERO registration number:** CRD42021279891

**Strengths and limitations of this study:**

- A living systematic review will allow continuous surveillance of emerging literature on different lifestyle interventions in women with a history of GDM and allow identification of effective strategies for diabetes prevention.
- We estimate considerable heterogeneity of interventions which may limit our ability to make clear conclusions.

## Introduction

Diabetes is fast becoming one of the biggest global health concerns with 463 million adults reported to be living with diabetes in 2019 [1]. Concerningly, further 1 in 2 adults are reported to have diabetes but are undiagnosed [1]. Diabetes is a non-communicable chronic disease with a progressive loss of β-cell mass and/or function that causes uncontrolled glucose levels in the blood [2]. It is thought to be caused by various genetic and environmental factors and can be classified into; Type-1 Diabetes Mellitus, Type-2 Diabetes Mellitus (T2DM), Gestational Diabetes Mellitus (GDM) and Diabetes caused by other causes [2]. Type 2 Diabetes is most prevalent and related to rising levels of obesity, unhealthy diets and physical inactivity [3]. Overwhelming studies and reviews show lifestyle interventions to be beneficial in reducing the prevalence of diabetes through weight loss and improvements in metabolic profile by making lifestyle changes such as diet and physical activity [4, 5], with further evidence of the efficacy of pharmacological interventions [6].

Gestational Diabetes Mellitus, previously considered a transient condition, is now an established risk factor for long-term diabetes. Women whose blood glucose levels do not return to normal soon after birth are at a particularly high risk of developing T2DM and consequent heart and blood vessel disease. While prevalence of GDM is markedly variable, it ranges between 2 – 25% globally [7]. In one longitudinal study, the risk of diabetes in women with GDM, compared to those without GDM, was estimated to be 17.9-fold [8]. The risk of developing T2DM appears to be particularly great in the first few years after delivery and at below 40 years of age [9]. This provides a compelling case for early prevention to reduce the burden of disease. However, having a baby presents new challenges to mothers and self-care is often not prioritised.

In general populations, lifestyle interventions have shown to be beneficial in reducing the prevalence of diabetes through weight loss and improved metabolic profile, and this has also been shown in women who had GDM. A recent systematic review of 15 Randomised Controlled Trials found that while postpartum lifestyle interventions were beneficial in 1733 participants, interventions implemented during pregnancy in 2883 participants were ineffective in preventing T2DM [10]. Interventions initiated within three years postpartum were also identified to be highly effective in reducing the risk of subsequent T2DM [10]. However, the review acknowledges several limitations, including the language barrier where they only included publications in English and Chinese, and notably the sample size of the included RCTs being relatively small.

## Methods

This study was performed according to the 2015 Preferred Reporting Items for Systematic reviews and Meta-analysis Protocols (PRISMA-P) statement.[11] The study protocol was registered with the International Database of Prospectively Registered Systematic Reviews in Health and Social care (PROSPERO) - CRD42021279891.

### Objective

This Living Systematic Review (LSR) aims to determine the effectiveness of postpartum interventions in women with a recent history of GDM in reducing the incidence of subsequent prediabetes and T2DM development.

### Eligibility criteria

Studied selected for inclusion will follow a pre-specified Population, Intervention, Comparison, Outcome, Timing and Study design (PICOTS) criteria.

#### Population

We will include studies in women within five years postpartum of a pregnancy complicated by gestational diabetes and who are currently not diagnosed with T2DM. GDM may be defined according to any recognised diagnostic criteria or based on medical record documentation.

#### Intervention

We will include studies that evaluate the effectiveness of any form of intervention for at least 12 months. Intervention may consist of lifestyle, behavioural and/or psychological changes as well as pharmacotherapy. These interventions may be standalone or in combination.

#### Comparison

We will include studies that have a comparison group, which may include usual care, placebo or another intervention.

#### Outcomes

##### Primary Outcome

Incidence of T2DM (defined as per standard criteria) and/or changes in glycaemic levels (fasting blood glucose level, oral glucose tolerance test and/or HbA1c), at the end of follow-up.

##### Secondary Outcomes

Changes to body weight, body mass index, waist circumference and other cardiometabolic biomarkers.

#### Timing of outcomes

We will include studies reporting baseline measurement of outcomes and at least one final measure at the end of the intervention period. We will include intervention of at least 12 months and include outcomes in any subsequent follow-up.

The living status of the systematic review will be maintained for five years after the protocol publication. The baseline living systematic review and prospective meta-analysis is planned to start from November 2021. An updated search will be conducted every 12 months.

### Study design

We will include any peer-reviewed RCT (individually randomised, cluster, stepped wedge, other) in any language. Pre-prints, theses, or dissertations without formal peer-review will not be included.

### Information source

Systematic research of online databases of PubMed, EMBASE and Web of Science will be undertaken. Two independent authors will search each database separately using the initial search strategy developed using PubMed and adapted as required for other databases. The reference lists of eligible articles will be screened for additional eligible articles. We will contact the corresponding authors of papers for any missing information.

### Search Strategy

A search strategy was developed using a combination of MeSH terms, keywords and their variations pertinent to our population, intervention and outcomes of interest, including ‘Gestational Diabetes Mellitus, ‘lifestyle’, ‘exercise’, ‘physical activity’, ‘diabetes mellitus’, ‘body weight’. The full search strategy is made available in Appendix A.

### Study records

#### Data management

Search results will be imported into reference management software Endnote X9. Duplicates will be removed, and individual search results will then be systematically screened for selection.

#### Selection process

Two reviewers will independently review the title and abstracts of studies derived from the search strategy against the predefined inclusion and exclusion criteria. After the initial screen, the full text of suitable studies will be screened for potentially relevant trials. Disagreements between study selection between two reviewers will be resolved by consensus involving a third reviewer. A screening process flowchart as per PRISMA recommendation will be constructed to summarise the process.

### Data extraction form

All the identified variables will be extracted and saved in a predefined data extraction form. Two independent authors will extract data, and any discrepancies will be resolved by consensus with a third author. The entire review team will review the data extraction form and pilot for the first three studies before commencing formal extraction.

### Assessment of risk of bias

Two reviewers will independently perform the risk of bias assessment for each published trial using the Cochrane risk-of-bias tool (RoB 2) [12]. Studies will be classified into ‘low risk of bias’, ‘some concern of bias’ and ‘high risk of bias’ using the Rob 2 tool by assessing several domains including random allocation sequence, allocation sequence concealment, blinding, outcome assessment, missing data, analysis methods. A risk-of-bias summary chart will be constructed using a ‘traffic light’ plot.

### Data Synthesis

A narrative summary of the effect of interventions categorized accordingly, in women with GDM on our primary outcome of interest, the incidence of T2DM, will be provided. Study characteristics will be presented in a table including country, design, intervention, comparator, outcomes, methods, funding and conflicts of interest.

We will perform Hartung-Knapp-Sidik-Jonkman random-effects aggregate data meta-analysis whenever it is feasible to do so (upon completion of the extraction process, we will perform a feasibility assessment to evaluate the appropriateness of pooling each outcome). For binary outcomes, we will use Risk Ratios (RR) and for continuous outcomes Mean Differences (MD): results will be presented in tabular format and by Forest Plots reporting also prediction intervals. We will assess quantitative heterogeneity by a formal test of homogeneity and evaluating the proportion of total variability due to heterogeneity rather than by sampling error (I^2^). Subgroup analyses will include but not limited to participant characteristics (BMI and ethnicity) and study characteristics (type of intervention, risk of bias, duration of follow-up and adherence to intervention). If appropriate to do so, we will also conduct univariable meta-regressions.

We will assess small-study effects by regression-based Egger test and eyeball evaluation of the contour-enhanced funnel plots.

Analyses will be based on reported data and intention to treat analyses: we will try to obtain missing outcome data from the original study authors. Complete case analyses will be performed as we will not impute any missing data. For the cluster randomized trials included we will make sure that their results take account of the cluster-effects: we will take account of clustering by adjusting the raw data for the design effect by using the effective sample size approach (i.e. the original sample size is divided by the design effect which is 1+(average cluster size-1)*Intracluster Correlation Coefficient [13]. We will perform statistical analyses using Stata 17 (StataCorp LLC, College Station, TX, USA) and/or R.

## Data Availability

The search strategy is in the appendix. Data extraction form is accessible upon
reasonable request.

### Appendix A. Search Strategy

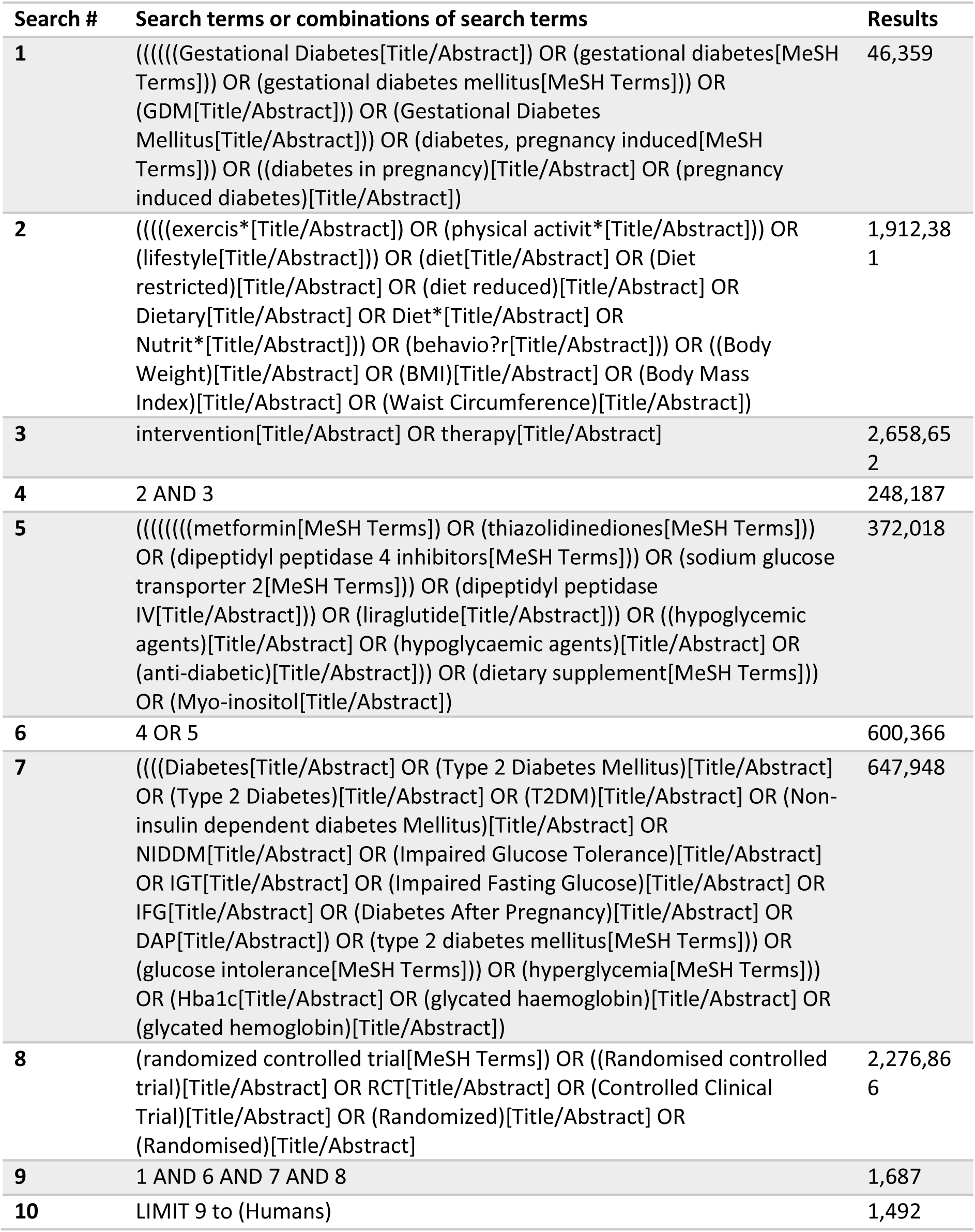

